# Skeletal Age for mapping the impact of fracture on mortality

**DOI:** 10.1101/2022.09.09.22279789

**Authors:** Thach Tran, Thao Ho-Le, Dana Bliuc, Bo Abrahamsen, Louise Hansen, Peter Vestergaard, Jacqueline R. Center, Tuan V. Nguyen

## Abstract

**Objectives:** to propose a novel “Skeletal Age” metric as the age of an individual’s skeleton resulting from a fragility fracture to convey the combined risk of fracture and fracture-associated mortality for an individual with specific risk profile.

**Design:** a retrospective population-based cohort study.

**Setting:** hospital records from the Danish National Hospital Discharge Register that includes the whole-country data of all contacts to health care system.

**Participants:** 1,667,339 adults in Denmark born on or before 1 January 1950, who were followed up to 31 December 2016 for incident low-trauma fracture and mortality.

**Main outcome measures:** fracture and chronic diseases recorded within 5 years prior to the index fracture were identified using ICD-10 codes. Death was ascertained from the Danish Register on Causes of Death. We used Cox’s proportional hazards regression to estimate the hazard ratio of mortality following a fracture, and then used the Gompertz law of mortality to transform the hazard ratio into life expectancy for a specific fracture site. The difference between life expectancy associated with a fracture and background population life expectancy is regarded as the years of life lost. Skeletal age is then operationally defined as an individual’s current age plus the years of life lost.

**Results:** during a median follow-up of 16.0 years, 95,372 men and 212,498 women sustained a fracture, followed by 41,017 and 81,727 deaths, respectively. A fracture was associated with 1 to 4 years of life lost dependent on fracture site, gender and age, with the greater loss being observed in younger men with a hip fracture. Hip, proximal and lower leg fractures, but not distal fractures, were associated with a substantial loss in life expectancy. A 60-year-old man with a hip fracture is expected to have a skeletal age of 66.1 years old (95% CI: 65.9, 66.2).

**Conclusion:** we propose to use skeletal age as a metric to assess fracture risk for an individual and thus improve doctor-patient risk communication.

**What have been known on this topic?:** Fragility fracture is associated with increased mortality risk, however it is currently underdiagnosed and undermanagement globally.

Despite the excess mortality after fracture, mortality is never a part of doctor-patient communication about treatment or risk assessment, due to a lack of an intuitive method of conveying risk as the traditional probability-based risk is counter-intuitive and hard to understand.

In engineering, “effective age” is the age of a structure based on its current conditions, and, in medicine, the effective age of an individual is the age of a typical healthy person who matches the specific risk profile of this individual.

**What this study adds:** We advanced the concept of “Skeletal Age” as the age of an individual’s skeleton resulting from a fragility fracture using data from a nationwide cohort of 1.7 million adults aged 50+ years old in Denmark.

Unlike the existing probability-based risk metrics, skeletal age combines the risk that an individual will sustain a fracture and the risk of mortality once a fracture has occurred, making the doctor-patient communication more intuitive and possibly more effective.

## INTRODUCTION

Fragility fracture is a direct consequence of osteoporosis, just as stroke is a consequence of hypertension. Fracture, especially hip fracture, is associated with an increased risk of mortality. Indeed, patients with a fragility fracture have on average a two-fold increase in the risk of mortality^1^. Between 22%^2^ and 58%^3^ of patients with a hip fracture die within 12 months post fracture. The identification of high-risk individuals for early intervention is a major priority in the control of osteoporotic fractures in the general community. In randomized controlled trials, treating high-risk individuals reduces the risk of fracture^4^ and the risk of mortality following a fracture^5^. However, the treatment uptake has been low, with only 40% of hip fracture patients in the United States being given osteoporosis treatment within 1 month after discharge in 2002, which was then halved in 2011^6^. This undertreatment is considered a crisis in the management of osteoporosis globally.

Despite the excess mortality after fracture, mortality is not part of doctor-patient communication about treatment or risk assessment, because there is a lack of an intuitive method of conveying risk. Traditionally, relative risk (e.g., “the risk of mortality is increased by 2-fold”) has been used as a metric of excess risk, but this metric often gives an exaggerated impression^7^, and is perceived as larger than the absolute risk^8^. On the other hand, absolute risk in terms of probability over a period of time is much harder to understand^9^, even for doctors and patients^10^. Thus, there is an urgent need for a more informative metric to internalize the combined risks of fracture and mortality in a way that patients and doctors can comprehend easily. The concept of “Effective Age” is useful here. In engineering, effective age is the age of a structure based on its current conditions, whereas in medicine, the effective age of an individual is the age of a typical healthy person who matches the specific risk profile of this individual^11^. Depending on whether the risk profile is not healthy (i.e., presence of risk factors) or healthy (i.e., absence of risk factors), the effective age is older or younger, respectively than the chronological age^11^. The best known effective age in medicine is “heart age” or “vascular age”^12^ and “lung age”^13^. The use of heart age and lung age has resulted in a better clinical impact than the traditional absolute risk metric^14^.

We consider that patients with osteoporosis and the general public are not sufficiently informed about the risk of post-fracture mortality, and this might have contributed to the current crisis of under management of osteoporosis^15 16^. In an effort to improve the management of osteoporosis, we advance the idea of “Skeletal Age” which is conceptually defined as the age of one’s skeleton as a consequence of fragility fracture. This new metric is not only one that quantifies the impact of fracture on mortality but also one that captures the risk of fracture and the risk of post-fracture mortality.

The aim of this study was to analyze the relationship between fracture and mortality, and then transform that relationship into the skeletal age for each individual fracture site using the Danish National Hospital Discharge Registry (NHDR). The NHDR data are ideal for this analysis because, apart from comorbidities at the individual patient level, it has documented the incidence of fractures and post-fracture mortality for the entire Danish population.

## MATERIALS AND METHODS

### Study design

The Danish National Hospital Discharge Registry can be viewed as a retrospective population-based cohort. This analysis included all adults aged 50 years old and older as at 1 January 2001 in Denmark whose health status had been followed up until 31 December 2016 for mortality. Individuals who had sustained a fracture at 45+ years old between 1996 and 2000 were excluded to avoid potential bias that the incident fracture analysed in this study was a second fracture (Figure 1). This is not a clinical trial. This analysis (Statistics Denmark project number 706667) was approved by the National Board of Health, the Danish Data Protection Agency, and Statistics Denmark, and subject to independent control and monitoring by The Danish Health Data Authority. Written informed consent is waived for routinely collected, pseudonymized registry data.

**Figure 1:**
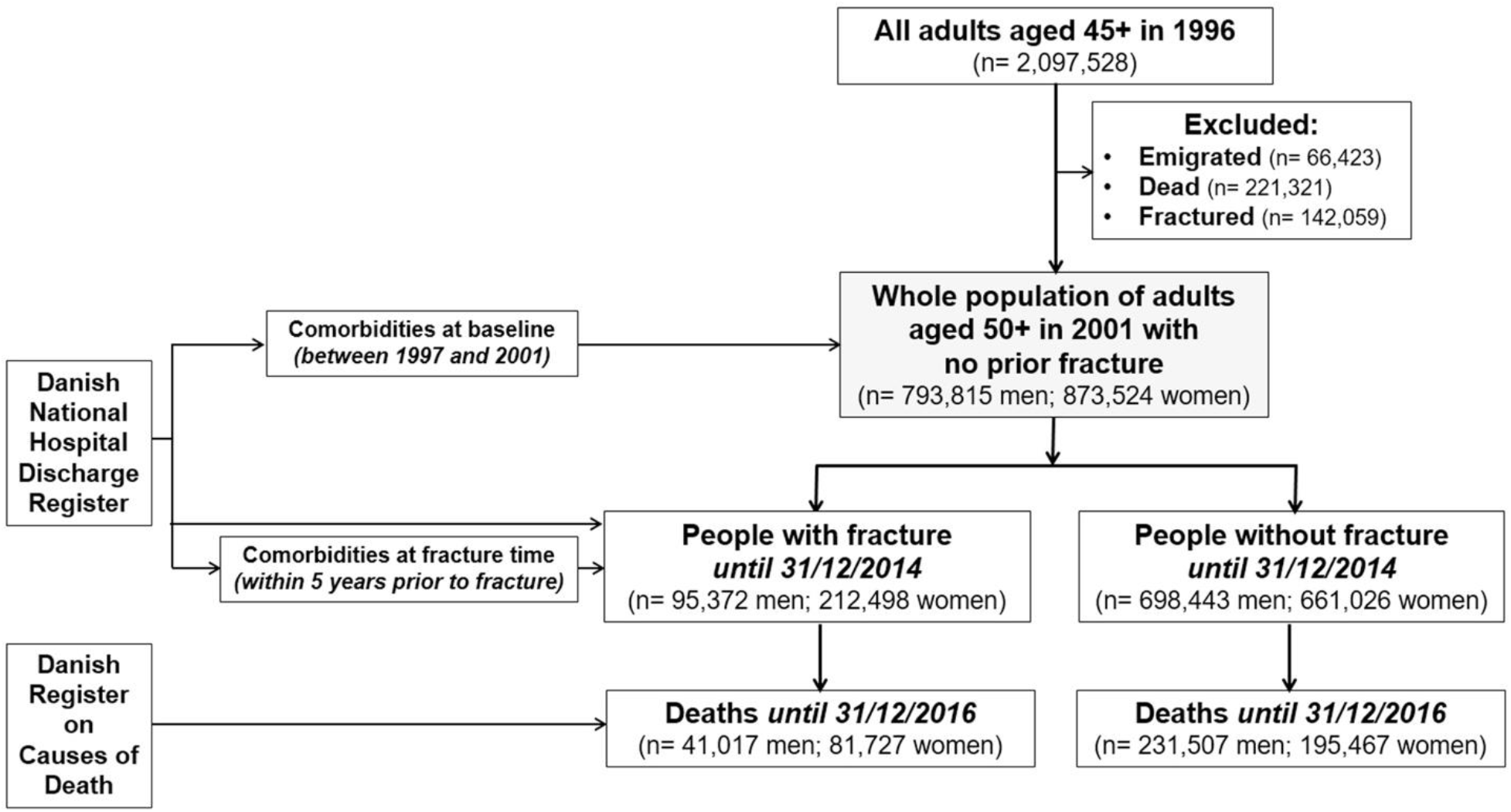
Flowchart of recruitment and follow up.

### Ascertainment of fracture and mortality

The initial incident fracture was defined as the first low-trauma fracture reported between 1 January 2001 and 31 December 2014. When more than one fracture occurred during a single event, only the fracture at the most proximal site was considered. We used the International Statistical Classification of Disease and Related Health Problems, tenth version (ICD-10) codes to identify individuals with specific fracture sites including hip, femur, pelvis, vertebrae, humerus, rib, clavicle (collectively known as proximal fractures), forearm, lower leg, knee, ankle, foot and hand (collectively known as distal fractures) from the Danish NHDR (Table S1). Face, skull, finger or toe fractures and high-trauma fractures due to traffic accidents were excluded.

The study participants were followed up to 31 December 2016 for mortality, allowing at least two years of follow up post fracture. Death was ascertained from the Danish Register on Causes of Death.

### Covariates assessment

The predefined covariates included age and comorbidities. We used the ICD-10 from the NHDR that includes any diagnosis documented between 1996 and 2000 to operationally define comorbidities at the study entry (i.e., 1 January 2001), and those within 5 years prior to the initial fracture to define comorbidities at fracture time. The severity of comorbidities was summarised using the Charlson index^17^.

### Statistical analysis

Skeletal age is operationally defined as the sum of the chronological age and the change in life expectancy associated with fracture. The changes in life expectancy resulted from a specific fracture were computed incorporating (i) the association between individual fracture site and mortality from a Cox’s proportional hazards regression and (ii) the baseline hazards described by Gompertz distribution and the population life expectancy from the national lifetable data^18^.

First, a Cox’s proportional hazards regression was used to quantify the association between a specific fracture and mortality, in which both fracture and confounding variables were analysed in a time-dependent manner. The models adjusted for age and severity of comorbidities^17^. Age at baseline and that at the time of fracture were also used. For individuals without fracture, the follow-up time was calculated from the study entry to the study end (31 December 2016) or date of death, whichever came first; while their covariates at baseline were adjusted in the analysis model (Figure S1). By contrast, the follow-up time for those with an incident fracture was split into the pre- and post-fracture period for which the clustering effect was accounted for in the Cox’s proportional hazards regression. The pre-fracture period was the time interval between the study entry and date of fracture, and used the covariates at study entry; whereas the post-fracture period was between the date of fracture and date of death or study end, whichever came first, and included the covariates at the time of fracture. The proportional hazards assumption was graphically checked using the Schoenfeld’s residuals.

Second, we used the Gompertz law of mortality and the Danish national lifetable data to transform the fracture-mortality association into life expectancy as a result of a fracture^18^. The Gompertz law of mortality indicates the annual risk of dying at the age *t* can be expressed as *h(t)* = *Be*^*kt*^ in which *B* ∼ 0.0000189 in women and 0.0000347 in men^19^. Under the Gompertz law of mortality, the annual risk of dying associated with ageing one year is remarkably consistent between ages 50 and 95 across ethnicities and over time^20 21^. Assuming the Gompertz baseline hazard, the log-hazards of mortality to specific fracture site *i* at the fracture time *t* is *a*+*bt*, where *a* and *b* are the estimated regression coefficients. The life expectancy for a specific fracture site at the fracture age z under the Gompertz distribution 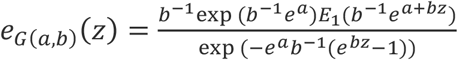,where *E*_1_(*b*^1^*e*^*a*+*bz*^) denotes the exponential integral. The loss of life years associated with specific fracture site was then calculated as the difference between the estimate life expectancy associated with fracture *e*_*G*(*a,b*)_*(z)* and the population life expectancy^18^. The skeletal age for each individual site of fracture was the sum of the individual’s chronological age at the time of fracture and the loss of life years as a result of the fracture. The 95% confidence interval of the skeletal age was computed using the confidence limits of the adjusted hazards ratios to account for uncertainty of the magnitude of association between individual site of fracture and mortality. The analyses were performed using Stata (*Stata Statistical Software: Release 16*. College Station, TX: StataCorp LLC) and the R statistical environment on a Windows platform^22^.

### Patient involvement

This study was an analysis of routinely collected, pseudonymized registry data. Neither were patients involved in generation of the research question or development of the outcome measures, nor the design and conduct of the study. The results will be disseminated to public through the public relations department of the University of Technology Sydney, Sydney, Australia.

## RESULTS

### Incidence of fractures

The present analysis was based on data from 793,815 men and 873,524 women aged 63.9 (standard deviation [SD] 10.3) and 65.5 (11.3) years old as at 1 January 2001 (Figure 1). During a median of 16 years of follow up (interquartile range [IQR]: 6.3, 16.0 years), 95,372 men and 212,498 women had sustained a fracture. The incidence of fractures was 10.9 fractures/1,000 person-years (95% confidence interval [CI]: 10.8, 11.0) in men and 23.2 fractures/1,000 person-years (23.1, 23.3) in women. Collectively, forearm, hip and humerus fractures accounted for 55% of all fractures in men and 70% in women.

As expected, men and women with a fracture were on average older than those without a fracture. Individuals with a fracture had more comorbidities than those who did not sustain a fracture (Table 1). For instance, the prevalence of myocardial infarction among patients with a fracture (9.3% in men and 4.7% in women) was higher than that among individuals without a fracture (4% in men and 1.5% in women). Similar trend was also observed for stroke, diabetes and cancer.

**Table 1:** Characteristics of the study population at baseline stratified by gender, fracture and mortality status.

| Characteristic | Non-fracture group |  | Fracture group |  |
| --- | --- | --- | --- | --- |
|  | Alive | Dead | Alive | Dead |
| **Men** |  |  |  |  |
| Number of individuals | 466936 | 231455 | 54486 | 40948 |
| Age at baseline<sup>1</sup> | 59.8 (7.7) | 71.1 (10.4) | 61.3 (8.9) | 72.4 (10.4) |
| Age at fracture<sup>1</sup> |  |  | 68.2 (9.9) | 77.8 (10.5) |
| Comorbidities* |  |  |  |  |
| *Charlson comorbidity index*<sup>2</sup> | 0 (0, 0) | 0 (0, 1) | 0 (0, 1) | 1 (0, 2) |
| 0 | 404415 (86.7%) | 140157 (60.6) | 32294 (59.4%) | 12553 (30.7%) |
| 1-2 | 56701 (12.1%) | 70904 (30.6%) | 17567 (32.3%) | 19011 (46.5%) |
| 3-4 | 3328 (0.7%) | 9203 (4.0%) | 2855 (5.3%) | 5209 (12.7%) |
| 5+ | 2492 (0.5%) | 11191 (4.8%) | 1639 (3.0%) | 4116 (10.1%) |
| *Specific comorbidities*<sup>3</sup> |  |  |  |  |
| Myocardial infarction | 18734 (4.0%) | 22243 (9.6%) | 5089 (9.3%) | 6041 (14.8%) |
| Congestive heart failure | 12814 (2.7%) | 26672 (11.5%) | 5108 (9.4%) | 8637 (21.1%) |
| Stroke | 21792 (4.7%) | 31317 (13.5%) | 9475 (17.4%) | 11538 (28.2%) |
| Peripheral vascular disease | 10912 (2.3%) | 17984 (7.8%) | 4263 (7.8%) | 5633 (13.8%) |
| Arrhythmias | 10835 (2.3%) | 18181 (7.9%) | 4885 (9.0%) | 7539 (18.4%) |
| Hypertension | 14340 (3.1%) | 15177 (6.6%) | 7712 (14.2%) | 7870 (19.2%) |
| Diabetes mellitus with no chronic complications | 19805 (4.2%) | 21166 (9.1%) | 6297 (11.6%) | 6287 (15.4%) |
| Diabetes mellitus with chronic complications | 6120 (1.3%) | 8205 (3.5%) | 2423 (4.5%) | 2744 (6.7%) |
| Renal disease | 4290 (0.9%) | 8754 (3.8%) | 2149 (3.9%) | 3604 (8.8%) |
| Cancer | 26126 (5.6%) | 43484 (18.8%) | 8938 (16.4%) | 11800 (28.8%) |
| Metastatic tumours | 1894 (0.4%) | 9478 (4.1%) | 1025 (1.9%) | 3015 (7.4%) |
| Dementia | 2869 (0.6%) | 7537 (3.3%) | 2690 (4.9%) | 6075 (14.8%) |
| Chronic lung disease | 16600 (3.6%) | 25597 (11.1%) | 6778 (12.4%) | 8673 (21.2%) |
| Rheumatologic | 4881 (1.1%) | 4569 (2.0%) | 1711 (3.1%) | 1610 (3.9%) |
| Mild liver disease | 2647 (0.6%) | 3749 (1.6%) | 1483 (2.7%) | 1788 (4.4%) |
| Moderate/severe liver disease | 589 (0.1%) | 1448 (0.6%) | 433 (0.8%) | 775 (1.9%) |
| Hemiplegia | 661 (0.1%) | 927 (0.4%) | 462 (0.9%) | 479 (1.2%) |
| HIV/AIDS | 108 (0.02%) | 81 (0.03%) | 37 (0.07%) | 23 (0.06%) |
| **Women** |  |  |  |  |
| Number of individuals | 465559 | 195404 | 130771 | 81727 |
| Age at baseline<sup>1</sup> | 60.5 (8.3) | 74.0 (11.1) | 63.9 (9.7) | 76.3 (9.9) |
| Age at fracture<sup>1</sup> |  |  | 70.9 (10.2) | 81.3 (9.6) |
| Comorbidities* |  |  |  |  |
| *Charlson comorbidity index*<sup>2</sup> | 0 (0, 0) | 0 (0, 1) | 0 (0, 1) | 1 (0, 2) |
| 0 | 400005 (85.9%) | 121773 (62.3%) | 81574 (62.4%) | 32093 (39.3%) |
| 1-2 | 59923 (12.9%) | 57569 (29.5%) | 41114 (31.4%) | 35866 (44.0%) |
| 3-4 | 2975 (0.6%) | 6320 (3.2%) | 5196 (4.0%) | 7960 (9.8%) |
| 5+ | 2656 (0.6%) | 9742 (5.0%) | 2887 (2.2%) | 5695 (7.0%) |
| *Specific comorbidities*<sup>3</sup> |  |  |  |  |
| Myocardial infarction | 7164 (1.5%) | 10885 (5.6%) | 6109 (4.7%) | 7562 (9.3%) |
| Congestive heart failure | 7307 (1.6%) | 18394 (9.4%) | 8056 (6.2%) | 13615 (16.7%) |
| Stroke | 16329 (3.5%) | 22759 (11.7%) | 17143 (13.1%) | 19247 (23.6%) |
| Peripheral vascular disease | 6517 (1.4%) | 10531 (5.4%) | 6196 (4.7%) | 7309 (9.0%) |
| Arrhythmias | 7671 (1.7%) | 13115 (6.7%) | 7947 (6.1%) | 11440 (14.0%) |
| Hypertension | 12910 (2.8%) | 12501 (6.4%) | 17970 (13.7%) | 15727 (19.3%) |
| Diabetes mellitus with no chronic complications | 14151 (3.0%) | 14020 (7.2%) | 9960 (7.6%) | 9214 (11.3%) |
| Diabetes mellitus with chronic complications | 3367 (0.7%) | 4461 (2.3%) | 2969 (2.3%) | 3126 (3.8%) |
| Renal disease | 2386 (0.5%) | 4217 (2.2%) | 2697 (2.1%) | 3561 (4.4%) |
| Cancer | 30913 (6.6%) | 34928 (17.9%) | 20246 (15.5%) | 18785 (23.0%) |
| Metastatic tumours | 2302 (0.5%) | 8872 (4.5%) | 2221 (1.7%) | 4632 (5.7%) |
| Dementia | 3035 (0.7%) | 7367 (3.8%) | 7169 (5.5%) | 13136 (16.1%) |
| Chronic lung disease | 18759 (4.0%) | 20946 (10.7%) | 15065 (11.5%) | 14018 (17.2%) |
| Rheumatologic | 10392 (2.2%) | 7064 (3.6%) | 7482 (5.7%) | 5256 (6.4%) |
| Mild liver disease | 2599 (0.6%) | 2271 (1.2%) | 2165 (1.7%) | 1802 (2.2%) |
| Moderate/severe liver disease | 296 (0.1%) | 686 (0.4%) | 422 (0.3%) | 643 (0.8%) |
| Hemiplegia | 524 (0.1%) | 573 (0.3%) | 577 (0.4%) | 475 (0.6%) |
| HIV/AIDS | 14 (0.0%) | 9 (0.0%) | 12 (0.01%) | 4 (0.0%) |

Notes: <sup>1</sup>: mean (SD); <sup>2</sup>: median (IQR); <sup>3</sup>: number (%). \*: at baseline for individuals without fracture or at fracture time for fracture patients. Comorbidities included not only chronic diseases that require hospitalisation, but also those documented as either secondary diagnoses or at outpatient or emergency visits.

### Incidence of mortality

During a median follow-up of 6.5 years (6.0 years (IQR: 2.5, 10.0) in men, 6.7 years (3.3, 10.8) in women), 272,524 men and 277,194 women had died, yielding mortality incidence rates of 6.55 deaths/100 person-years (95% CI: 6.48, 6.61) in men and 5.42 deaths/100 person-years (5.39, 5.46) in women. More importantly, the rate of mortality among patients with a fracture (5.4 per 100 person-years in men and 6.5 per 100 person-years in women) was greater than among those without a fracture (2.6 per 100 person-years in men and 2.2 per 100 person-years in women). Analysis by fracture site revealed that men and women with a fracture at the hip, pelvis or vertebra had a much greater risk of mortality than other fractures (Table 2).

**Table 2:** Incidence of mortality following specific fracture sites stratified by gender.

| Fracture | Number | Age at fracture (years) | Number of deaths | Follow-up time (person-years) | Rate* of mortality (95% CI) |
| --- | --- | --- | --- | --- | --- |
| **Men** |  |  |  |  |  |
| No fracture | 698391 | 63.6** (10.2) | 231455 | 9049194 | 2.6% (2.5, 2.6) |
| Any fracture | 95434 | 72.3 (11.2) | 81689 | 1506940 | 5.4% (5.4, 5.5) |
| Hip fracture | 25700 | 79.5 (9.7) | 16864 | 107789 | 15.6% (15.4, 15.9) |
| Femur | 1908 | 74.5 (10.8) | 993 | 10468 | 9.5% (8.9, 10.1) |
| Pelvis | 1302 | 77.0 (10.9) | 748 | 6465 | 11.6% (10.8, 12.4) |
| Vertebrae | 6926 | 72.7 (10.5) | 3120 | 41746 | 7.5% (7.2, 7.7) |
| Humerus | 10132 | 72.6 (10.7) | 4818 | 62350 | 7.7% (7.5, 7.9) |
| Rib | 6874 | 69.7 (10.4) | 2346 | 50186 | 4.7% (4.5, 4.9) |
| Clavicle | 5210 | 68.2 (10.5) | 1633 | 39731 | 4.1% (3.9, 4.3) |
| Lower leg | 6304 | 67.0 (9.5) | 1794 | 52915 | 3.4% (3.2, 3.6) |
| Forearm | 15284 | 69.4 (10.2) | 4667 | 120719 | 3.9% (3.8, 4.0) |
| Knee | 1386 | 70.2 (10.1) | 447 | 10886 | 4.1% (3.7, 4.5) |
| Ankle | 1084 | 67.6 (9.8) | 273 | 9081 | 3.0% (2.7, 3.4) |
| Hand | 8320 | 67.7 (10.1) | 2176 | 70263 | 3.1% (3.0, 3.2) |
| Foot | 5004 | 65.3 (9.7) | 1069 | 44136 | 2.4% (2.3, 2.6) |
| **Women** |  |  |  |  |  |
| No fracture | 660963 | 64.5** (11.1) | 195404 | 8723863 | 2.2% (2.2, 2.3) |
| Any fracture | 212601 | 74.9 (11.2) | 40948 | 626733 | 6.5% (6.5, 6.6) |
| Hip fracture | 51674 | 82.1 (9.2) | 31801 | 259087 | 12.3% (12.1, 12.4) |
| Femur | 3526 | 78.9 (10.9) | 1919 | 19322 | 9.9% (9.5, 10.4) |
| Pelvis | 4923 | 81.2 (10.0) | 2891 | 26761 | 10.8% (10.4, 11.2) |
| Vertebrae | 9741 | 76.6 (10.6) | 4656 | 60295 | 7.7% (7.5, 7.9) |
| Humerus | 28317 | 74.4 (10.5) | 10325 | 202662 | 5.1% (5.0, 5.2) |
| Rib | 3158 | 74.1 (11.9) | 1215 | 22285 | 5.5% (5.1, 5.8) |
| Clavicle | 4465 | 72.7 (11.5) | 1583 | 31755 | 5.0% (4.7, 5.2) |
| Lower leg | 11469 | 70.1 (10.8) | 3131 | 94308 | 3.3% (3.2, 3.4) |
| Forearm | 68376 | 72.0 (10.3) | 18459 | 559406 | 3.3% (3.2, 3.3) |
| Knee | 2611 | 71.3 (9.7) | 630 | 21086 | 3.0% (2.8, 3.2) |
| Ankle | 1936 | 70.0 (10.6) | 496 | 15726 | 3.2% (2.9, 3.4) |
| Hand | 12489 | 69.6 (10.2) | 2682 | 108112 | 2.5% (2.4, 2.6) |
| Foot | 9916 | 67.8 (9.6) | 1901 | 86136 | 2.2% (2.1, 2.3) |

Notes: \*: rates were calculated as number of deaths/100 person-years; \*\*: age at baseline (years).

However, the above observation could be confounded by age and comorbidities, because individuals with a fracture were on average older and had more comorbidities than those without a fracture. Therefore, we employed Cox’s proportional hazards model to estimate the strength of association between fracture and mortality, adjusting for age and comorbidities (Figure 2). Individuals with any fragility fracture was associated with 30-45% increased hazard of death (adjusted hazard ratio = 1.46 (95% CI: 1.45, 1.48) in men; 1.28 (1.27, 1.30) in women). For the same age and comorbidity profile, men and women with a hip or femur fracture had almost 2-fold greater risk of death than those without a fracture.

**Figure 2:**
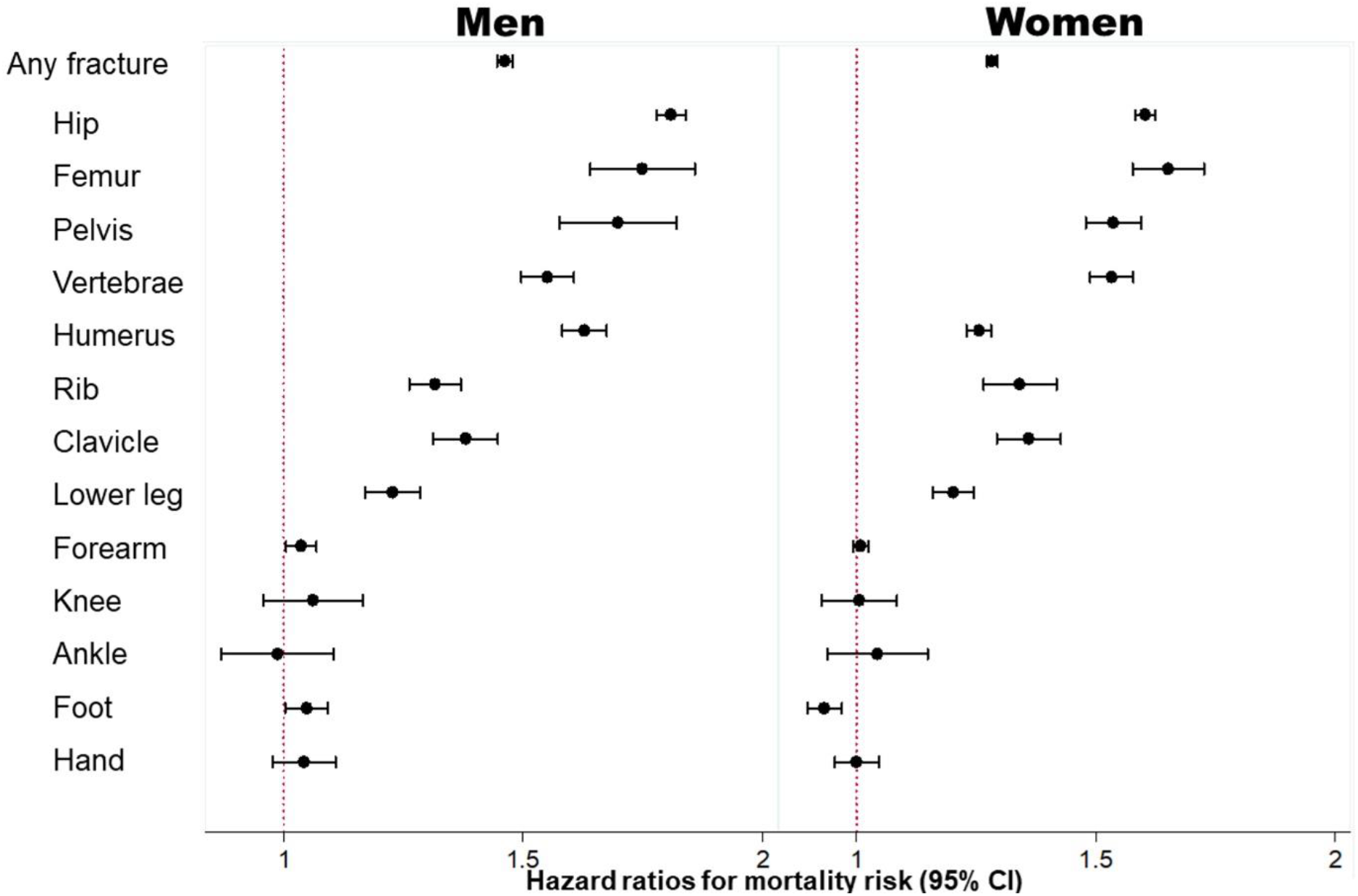
Association between specific fracture and mortality risk, adjusted for age and presence of comorbidities: hazard ratio and 95% confidence interval.

The increased risk of mortality was also observed among those with a proximal fracture, such as pelvis, vertebrae, humerus, rib, clavicle, and lower leg fracture after controlling for age and comorbidities. However, there was no significantly increased risk of deaths following a forearm, knee or foot fracture (Figure 2).

### Skeletal age

Based on the association between fracture and mortality and using the Gompertz law (see Methods), we estimated the skeletal age for each bone fracture and each chronological age (from the age of 50) in men and women (Figure 3, Table S2). Patients with a fracture -- any fracture at all -- have lost years of life, and hence their skeletal age was greater than their chronological age. For a given age, men with a fracture on average had greater skeletal age than women by approximately 1 year (54.2 vs 53.4 years). In either sex, the loss of years of life was more pronounced in younger age groups and gradually converged in the older age groups.

**Figure 3:**
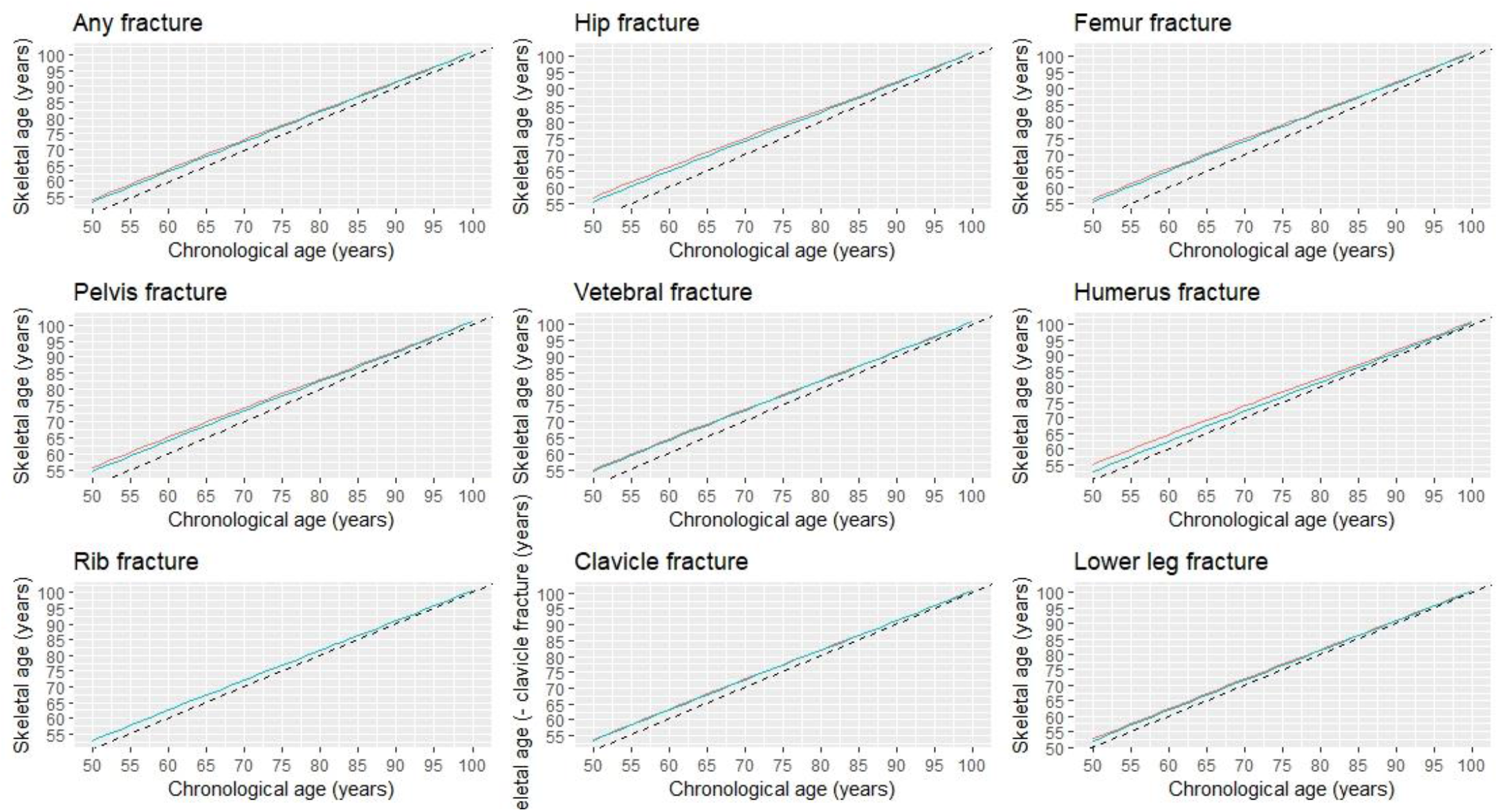
Skeletal age by specific fracture site and chronological age at fracture. **Legend**: dashed lines indicate perfect concordance between the chronological and skeletal ages; solid lines indicate the differences between the chronological age at the time of fracture and corresponding skeletal age for men (red) and women (blue).

As expected, patients with a hip fracture had the highest skeletal age than those with other fractures. For example, a 70-year-old man who had sustained a hip fracture would have a skeletal age of 75 years (e.g., a loss of 5 years of life); however, if a 50-year-old man with a hip fracture would have a skeletal age of 56.8 years which is equivalent to almost 7 years of life lost. Other fractures such as femur, pelvis, vertebrae, and humerus also signified a significant loss of years of life (around 5 years); thus, 50-year-old patients with one of these fractures are estimated to have a skeletal age of around 55 years.

However, fractures are the rib, clavicle and lower leg were associated with lower years of life lost, and the skeletal age of patients with one of these fractures was generally lower than patients with a more serious fracture (e.g., hip fracture). For instance, a 60-year-old patient with a lower leg fracture would be expected to have a skeletal age of 62.4 years for men or 61.9 years for women (Table S3).

## DISCUSSION

It has been well established that fracture, especially hip fracture, is associated with an increased risk of mortality, and this excess risk is commonly expressed in terms of the relative risk metric. Here, we proposed a new effective age metric called “Skeletal Age” to quantify the impact of fracture on mortality. Using data from a nationwide cohort of 1.7 million adults aged 50+ years old in Denmark, we showed that almost all types of fracture were associated with a loss of years of life, indicating that the skeletal age of individuals suffering from a fracture is greater than their chronological age. This finding has important implications that we discuss below.

### Findings in context

Our finding confirmed the previous studies^23-28^ that patients with a fragility fracture were associated with a significantly greater risk of mortality than their similarly aged and gender counterparts without fracture who had the same comorbidity profile. Our finding is also consistent with an earlier Danish study demonstrating reduced life expectancy in patients, with or without fractures, at the time of beginning osteoporosis treatment^29^. However, the magnitude of the association between individual sites of fracture observed in our study is slightly lower than that documented in several previous cohort studies^24-28^, with hip fractures being associated with 1.7-to-6-fold increased risk of death^27^. This discrepancy could be due to the difference in study populations. While cohort studies usually recruit healthy participants, our national registry-based study was able to document all individuals in Denmark. As a result, our control group (i.e., those without a fracture) included individuals in poor health conditions with multiple comorbidities who are usually unable to be recruited in a cohort study.

Whether the association between fracture and mortality is causal or not is a matter of contention. It is commonly assumed that the association is confounded by comorbidities, but multiple studies have found that comorbidities contributed little to the excess risk of mortality among fractured patients^30 31^. In the present analysis we have also adjusted for comorbidities, and the fracture - mortality remained unchanged, suggesting that the association is unlikely due to comorbidities. The Study of Osteoporotic Fracture^32^ found that women who died after a vertebral fracture often had chronic obstructive pulmonary disease and pneumonia. Logically, if fracture is a causal factor that increases mortality risk, then treatment of patients with a fracture should reduce mortality risk, and that has been observed in a randomized controlled trial. Indeed, Lyles and colleagues showed that osteoporotic patients on zoledronic acid had a 28 per cent lower risk of all-cause mortality compared to those on placebo^33^, which was confirmed by a subsequent re-analysis^34^ that adjusted for baseline fracture risk. The beneficial effect of zoledronic acid on mortality was further observed in a recent study, in which osteopenic patients with a fracture on zoledronic acid had a lower risk of mortality (average relative risk reduction of 35 per cent) than those on placebo, although the difference did not reach the conventional statistical significance.

Regardless of the nature of the association, mortality is not a component of doctor-patient communication. Existing fracture risk assessment models such as Garvan^35^ and FRAX^36^ provide the probability (i.e., absolute risk) of fracture over a 10-year period without any information on mortality. However, doctors and patients have difficulty in understanding and interpreting probability^10^. Only a fifth of a sample of highly educated American adults could understand one in 1000 is equivalent to 0.01%^37^. Conveying a low, though clinically significant absolute risk (e.g., “*Your risk of hip fracture over the next 10 years is 5%*”) might provide a false peace of mind and underestimated risk perception, leading to increasing possibilities of refusing the recommended treatment^7^. Thus, poor communication about fracture risk and mortality consequence might have contributed to the global crisis of undertreatment of osteoporosis.

### Potential implications

Based on the concept of effective age, we proposed the idea of Skeletal Age as a new metric for communicating the risk of mortality following a fracture. Unlike relative risk and absolute risk metrics which are based on probability, skeletal age is a natural frequency metric that has been consistently shown to be easier and more friendly to doctors and patients^8 10 38^. It is not straightforward to appreciate the importance of the two-fold increased risk of death (i.e., relative risk = 2.0) without knowing the background risk (i.e., 2 folds of 1% would remarkably differ from 2 folds of 10%). By contrast, for the same 2-fold mortality risk of hip fracture, telling a 60-year man with a hip fracture that his skeletal age would be 66 years old, equivalent to a 6-year loss of life, is more intuitive. In addition, the skeletal age can be also used to convey the possible benefit of treatment, providing an alternative metric to the conventional metrics such as relative risk or relative risk reduction. For instance, a patient might find the statement “*Zoledronic acid treatment helps a patient with a hip fracture gain 3 years of life*” much easier to understand and probably more persuasive than “*Zoledronic acid treatment reduced the risk of death by 28%*”^33^.

Skeletal Age is an addition to the already available metrics such as “Heart Age”, “Lung Age”, and more recently “Covid Age” that have been demonstrated to be superior to the conventional risk metrics^39-43^ in terms of positive behavioral changes. For instance, compared with usual care, individuals who were given heart age had a greater smoking cessation rate^41 42^, substantial improvement in weight, body mass index and physical activity^41^ and greater proportion of high-risk patients returning for a follow-up appointment^40^. The use of heart age also led to significantly greater improvement at 12 months in metabolic parameters^41^ or lipid profiles^39^. A recent cluster randomised controlled trial in Norway also found that heart age was a good way to communicate cardiovascular risk and to motivate individuals to reduce cardiovascular risk factors^43^. Similarly, the use of lung age could also lead to a clinically significant increase in the reported smoking cessation rates^44 45 46^. Collectively, these data suggest that effective age metrics can help patients better understand their risk and lead to preventive changes.

### Strengths and limitations of the study

Our findings should be interpreted within the context of their strengths and limitations. First, we included a whole-country population with long follow up and robust diagnostic data, minimizing potential selection bias and misclassification^47^, and allowing us to examine mortality risk following individual fracture sites. Second, the current study analysed both fracture and covariates, such as aging and the presence of comorbidities in a time-dependent manner, making it statistically rigorous in minimizing an immortal bias and sufficiently accounting for confounding effects. The analysis thus provided accurate estimates of the association between specific fracture sites and mortality risk as it was able to control for confounding effects at both the study entry and the time of fracture.

However, the use of registry-based data, originally documented and coded for administrative or reimbursement purposes is prone to variable data accuracy, the lack of specific clinical information and non-medical factors^48^. Fortunately, all essential study variables (i.e., comorbidities, fracture and death) were systematically obtained from the Danish NHDR that includes excellent, complete medical records and precise diagnoses for all individuals living in Denmark since 1995^49 50^. The use of diagnosis-based comorbidities, though possibly associated with lower sensitivity than the self-reported ones^51^ is able to capture the severe health disorders that prompt the participants to seek for medical assistance. Secondly, the analyses could not make adjustments for lifestyle factors and physical activity, which are usually obtained in a cohort study. As they are closely related to aging and the presence of comorbidities which were already accounted for in our analysis, making further adjustments for these lifestyle factors is unlikely to substantially modify the findings.

## Conclusions

We advanced the concept of skeletal age as the age of an individual’s skeleton resulting from a fragility fracture. Unlike existing metrics (e.g., relative risk, probability of fracture), skeletal age combines the risk that an individual will sustain a fracture and the risk of mortality once a fracture has occurred, making the doctor-patient communication more intuitive and possibly more effective. Given the evidence of successful implication of similar effective age metrics, skeletal age is expected to improve risk communication and ultimately improve treatment uptake among patients who are indicated for treatment. A randomized controlled trial aiming to compare the use of skeletal age and the current metrics in fracture risk communication is warranted.

## Data Availability

Patient level data cannot be shared without approval from data custodians owing to local information governance and data protection regulations.

## Authors’ roles

Tuan V. Nguyen, Thach Tran, and Thao Ho-Le contributed to study conceptualisation and design. Thach Tran, Thao Ho-Le, and Tuan V. Nguyen contributed to data analysis. Thach Tran, Thao Ho-Le, Dana Bliuc, Bo Abrahamsen, Louise Hansen, Peter Vestergaard, Jacqueline R. Center, and Tuan V. Nguyen contributed to data acquisition and interpretation. Thach Tran and Tuan V. Nguyen wrote the first draft. All authors contributed to revising and approving the final version of the manuscript.

## Ethical approval

This analysis (Statistics Denmark project number 706667) was approved by the National Board of Health, the Danish Data Protection Agency, and Statistics Denmark, and subject to independent control and monitoring by The Danish Health Data Authority. Written informed consent is waived for routinely collected, pseudonymized registry data.

## Transparency

The lead authors (Thach Tran and Tuan V. Nguyen) affirm that this manuscript is an honest, accurate, and transparent account for the study being reported; that no important aspects of the study have been omitted; and that any discrepancies from the study as originally planned have been explained.

## Dissemination to participants and related patient and public communities

We will disseminate a lay summary of our findings through the press release and other social media accounts.

## Appendix

**Table S1:** List of ICD-10 codes used to define specific fractures and comorbidities.

| Conditions | ICD-10 codes |
| --- | --- |
| **Fractures:** |  |
| *Proximal fractures:* |  |
| Hip | S72.0- S72.2 |
| Femur | S72.3-S72.9 |
| Pelvis | S32.3-S32.5 |
| Vertebrae | S22.0, S22.1, S32.0, S32.2, S32.7, S32.8, T08.x |
| Humerus | S42.x |
| Rib | S22.3-S22.4 |
| Clavicle | S42.0 |
| *Distal fractures:* |  |
| Forearm | S52.x |
| Lower leg | S82.2-S82.8 |
| Knee | S82.0 |
| Ankle | S82.5-S82.6 |
| Foot | S92.0-S92.3, S92.7, S92.9 |
| Hand | S62.0-S62.4, S62.8 |
| **Comorbidities:** |  |
| Myocardial infarct | I21.x, I22.x, I25.2 |
| Congestive heart failure | I09.9, I11.0, I13.0, I13.2, I25.5, I42.0, I42.5 - I42.9, I43.x, I50.x, P29.0 |
| Peripheral vascular disease | I70.x, I71.x, I73.1, I73.8, I73.9, I77.1, I79.0, I79.2, K55.1, K55.8, K55.9, Z95.8, Z95.9 |
| Cerebrovascular disease | G45.x, G46.x, H34.0, I60.x - I69.x |
| Cardiac valvular disease | A52.0, I05.x - I08.x, I09.1, I09.8, I34.x - I39.x, Q23.0 - Q23.3, Z95.2 - Z95.4 |
| Cardiac arrhythmias | I44.1 - I44.3, I45.6, I45.9, I47.x - I49.x, R00.0, R00.1, R00.8, T82.1, Z45.0, Z95.0 |
| Diabetes without chronic complication | E10.0, E10.1, E10.6, E10.8, E10.9, E11.0, E11.1, E11.6, E11.8, E11.9, E12.0, E12.1, E12.6, E12.8, E12.9, E13.0, E13.1, E13.6, E13.8, E13.9, E14.0, E14.1, E14.6, E14.8, E14.9 |
| Diabetes with chronic complication | E10.2 - E10.5, E10.7, E11.2 - E11.5, E11.7, E12.2 - E12.5, E12.7, E13.2 - E13.5, E13.7, E14.2 - E14.5, E14.7 |
| Any malignancy, including lymphoma and leukaemia, except malignant neoplasm of skin | C00.x - C26.x, C30.x - C34.x, C37.x - C41.x, C43.x, C45.x - C58.x, C60.x - C76.x, C81.x - C85.x, C88.x, C90.x - C97.x |
| Metastatic solid tumour | C77.x - C80.x |
| Rheumatic/Rheumatoid arthritis or collagen vascular disease | L94.0, L94.1, L94.3, M05.x, M06.x, M31.5, M32.x - M34.x, M08.x, M12.0, M12.3, M30.x, M31.0 - M31.3, M32.x - M35.x, M36.0, M45.x, M46.1, M46.8, M46.9 |
| Mild liver disease | B18.x, K70.0 - K70.3, K70.9, K71.3 - K71.5, K71.7, K73.x, K74.x, K76.0, K76.2 - K76.4, K76.8, K76.9, Z94.4 |
| Moderate or severe liver disease | I85.0, I85.9, I86.4, I98.2, K70.4, K71.1, K72.1, K72.9, K76.5, K76.6, K76.7 |
| Hypertension | I10.x, I11.x - I13.x, I15.x |
| Chronic pulmonary disease | I27.8, I27.9, J40.x - J47.x, J60.x - J67.x, J68.4, J70.1, J70.3 |
| Pulmonary circulation disorders | I26.x, I27.x, I28.0, I28.8, I28.9 |
| Dementia | F00.x - F03.x, F05.1, G30.x, G31.1 |
| Psychoses | F20.x, F22.x - F25.x, F28.x, F29.x, F30.2, F31.2, F31.5 |
| Depression | F20.4, F31.3 - F31.5, F32.x, F33.x, F34.1, F41.2, F43.2 |
| Neurological disorders | G10.x - G13.x, G20.x - G22.x, G25.4, G25.5, G31.2, G31.8, G31.9, G32.x, G35.x - G37.x, G40.x, G41.x, G93.1, G93.4, R47.0, R56.x |
| Hemiplegia or paraplegia | G04.1, G11.4, G80.1, G80.2, G81.x, G82.x, G83.0 - G83.4, G83.9 |
| Peptic ulcer disease | K25.x - K28.x |
| Chronic kidney disease | I12.0, I13.1, N03.2 - N03.7, N05.2 - N05.7, N18.x, N19.x, N25.0, Z49.0 - Z49.2, Z94.0, Z99.2 |
| Hypothyroidism | E00.x - E03.x, E89.0 |
| Coagulopathy | D65 - D68.x, D69.1, D69.3 - D69.6 |
| Obesity | E66.x |
| Weight loss | E40.x - E46.x, R63.4, R64 |
| Fluid and electrolyte disorders | E22.2, E86.x, E87.x |
| Anaemia | D50.0, D50.8, D50.9, D51.x - D53.x |
| Alcohol abuse | F10, E52, G62.1, I42.6, K29.2, K70.0, K70.3, K70.9, T51.x, Z50.2, Z71.4, Z72.1 |
| Drug abuse | F11.x - F16.x, F18.x, F19.x, Z71.5, Z72.2 |
| AIDS/HIV | B20.x - B22.x, B24.x |

**Table S2:** Skeletal age by specific fracture site and chronological age at fracture.

| **Chronological**<br>**age (years)** | **Any**<br>**fracture** | **Hip**<br>**fracture** | **Femur**<br>**fracture** | **Pelvis**<br>**fracture** | **Vertebral**<br>**fracture** | **Humerus**<br>**fracture** | **Rib**<br>**fracture** | **Clavicle**<br>**fracture** | **Lower**<br>**leg**<br>**fracture** |
| --- | --- | --- | --- | --- | --- | --- | --- | --- | --- |
| **Men** |  |  |  |  |  |  |  |  |  |
| 50 | 54.2 | 56.8 | 56.3 | 55.7 | 54.9 | 55.0 | 52.8 | 53.6 | 52.7 |
| 51 | 55.2 | 57.8 | 57.3 | 56.7 | 56.0 | 56.0 | 53.9 | 54.6 | 53.7 |
| 52 | 56.2 | 58.8 | 58.3 | 57.7 | 56.9 | 57.0 | 54.9 | 55.6 | 54.7 |
| 53 | 57.2 | 59.7 | 59.2 | 58.7 | 57.9 | 58.0 | 55.9 | 56.6 | 55.7 |
| 54 | 58.1 | 60.6 | 60.1 | 59.6 | 58.8 | 58.9 | 56.8 | 57.5 | 56.6 |
| 55 | 59.1 | 61.5 | 61.1 | 60.5 | 59.8 | 59.8 | 57.8 | 58.5 | 57.6 |
| 56 | 60.0 | 62.5 | 62 | 61.5 | 60.8 | 60.8 | 58.8 | 59.5 | 58.6 |
| 57 | 61.0 | 63.4 | 62.9 | 62.4 | 61.7 | 61.7 | 59.7 | 60.5 | 59.6 |
| 58 | 61.9 | 64.3 | 63.9 | 63.3 | 62.6 | 62.7 | 60.7 | 61.4 | 60.5 |
| 59 | 62.9 | 65.2 | 64.8 | 64.2 | 63.6 | 63.6 | 61.6 | 62.4 | 61.5 |
| 60 | 63.8 | 66.1 | 65.7 | 65.2 | 64.5 | 64.5 | 62.6 | 63.3 | 62.4 |
| 61 | 64.8 | 67.1 | 66.6 | 66.1 | 65.5 | 65.5 | 63.6 | 64.3 | 63.5 |
| 62 | 65.7 | 67.9 | 67.5 | 67.0 | 66.4 | 66.4 | 64.6 | 65.2 | 64.4 |
| 63 | 66.6 | 68.8 | 68.4 | 67.9 | 67.3 | 67.3 | 65.5 | 66.2 | 65.3 |
| 64 | 67.6 | 69.7 | 69.4 | 68.9 | 68.2 | 68.3 | 66.5 | 67.1 | 66.3 |
| 65 | 68.6 | 70.7 | 70.3 | 69.8 | 69.2 | 69.2 | 67.5 | 68.1 | 67.3 |
| 66 | 69.5 | 71.5 | 71.1 | 70.7 | 70.1 | 70.1 | 68.4 | 69.0 | 68.2 |
| 67 | 70.4 | 72.4 | 72.0 | 71.6 | 71.0 | 71.0 | 69.3 | 69.9 | 69.2 |
| 68 | 71.4 | 73.3 | 73.0 | 72.5 | 72.0 | 72.0 | 70.3 | 71.0 | 70.2 |
| 69 | 72.3 | 74.2 | 73.8 | 73.4 | 72.9 | 72.9 | 71.3 | 71.9 | 71.1 |
| 70 | 73.2 | 75.0 | 74.7 | 74.3 | 73.7 | 73.8 | 72.2 | 72.8 | 72.1 |
| 71 | 74.1 | 75.8 | 75.5 | 75.1 | 74.6 | 74.6 | 73.1 | 73.7 | 73.0 |
| 72 | 75.1 | 76.8 | 76.5 | 76.1 | 75.6 | 75.6 | 74.1 | 74.7 | 74.0 |
| 73 | 75.9 | 77.6 | 77.3 | 76.9 | 76.4 | 76.4 | 75.0 | 75.5 | 74.9 |
| 74 | 76.9 | 78.5 | 78.2 | 77.8 | 77.3 | 77.4 | 76.0 | 76.5 | 75.9 |
| 75 | 77.8 | 79.3 | 79.0 | 78.7 | 78.2 | 78.3 | 76.9 | 77.4 | 76.8 |
| 76 | 78.7 | 80.2 | 80.0 | 79.6 | 79.2 | 79.2 | 77.9 | 78.4 | 77.8 |
| 77 | 79.6 | 81.1 | 80.8 | 80.5 | 80.1 | 80.1 | 78.8 | 79.3 | 78.7 |
| 78 | 80.5 | 81.9 | 81.6 | 81.3 | 80.9 | 81.0 | 79.8 | 80.2 | 79.7 |
| 79 | 81.5 | 82.8 | 82.6 | 82.3 | 81.9 | 81.9 | 80.8 | 81.2 | 80.6 |
| 80 | 82.3 | 83.6 | 83.3 | 83.1 | 82.7 | 82.7 | 81.6 | 82.0 | 81.5 |
| 81 | 83.3 | 84.5 | 84.3 | 84 | 83.7 | 83.7 | 82.6 | 83.0 | 82.5 |
| 82 | 84.2 | 85.3 | 85.1 | 84.9 | 84.5 | 84.6 | 83.6 | 83.9 | 83.5 |
| 83 | 85.0 | 86.1 | 85.9 | 85.7 | 85.4 | 85.4 | 84.4 | 84.8 | 84.3 |
| 84 | 86.0 | 87.0 | 86.8 | 86.6 | 86.3 | 86.3 | 85.4 | 85.7 | 85.3 |
| 85 | 86.9 | 87.8 | 87.7 | 87.5 | 87.2 | 87.2 | 86.3 | 86.7 | 86.3 |
| 86 | 87.8 | 88.7 | 88.6 | 88.4 | 88.1 | 88.1 | 87.3 | 87.6 | 87.2 |
| 87 | 88.7 | 89.5 | 89.4 | 89.2 | 88.9 | 89.0 | 88.2 | 88.5 | 88.1 |
| 88 | 89.6 | 90.4 | 90.3 | 90.1 | 89.9 | 89.9 | 89.1 | 89.4 | 89.1 |
| 89 | 90.6 | 91.3 | 91.2 | 91.1 | 90.8 | 90.8 | 90.2 | 90.4 | 90.1 |
| 90 | 91.5 | 92.2 | 92.1 | 92.0 | 91.7 | 91.8 | 91.1 | 91.4 | 91.0 |
| 91 | 92.4 | 93.0 | 92.9 | 92.8 | 92.6 | 92.6 | 92.0 | 92.2 | 91.9 |
| 92 | 93.4 | 94.0 | 93.9 | 93.8 | 93.6 | 93.6 | 93.0 | 93.2 | 93.0 |
| 93 | 94.3 | 94.8 | 94.7 | 94.6 | 94.4 | 94.4 | 93.9 | 94.1 | 93.8 |
| 94 | 95.2 | 95.7 | 95.7 | 95.5 | 95.4 | 95.4 | 94.9 | 95.1 | 94.8 |
| 95 | 96.2 | 96.7 | 96.6 | 96.5 | 96.3 | 96.3 | 95.9 | 96.0 | 95.8 |
| 96 | 97.1 | 97.6 | 97.5 | 97.4 | 97.3 | 97.3 | 96.8 | 97.0 | 96.8 |
| 97 | 98.0 | 98.5 | 98.4 | 98.3 | 98.2 | 98.2 | 97.8 | 97.9 | 97.7 |
| 98 | 99.0 | 99.4 | 99.4 | 99.3 | 99.1 | 99.1 | 98.7 | 98.9 | 98.6 |
| 99 | 100.0 | 100.5 | 100.4 | 100.3 | 100.1 | 100.2 | 99.7 | 99.9 | 99.7 |
| 100 | 100.9 | 101.4 | 101.3 | 101.2 | 101.1 | 101.1 | 100.7 | 100.8 | 100.6 |
| **Women** |  |  |  |  |  |  |  |  |  |
| 50 | 53.4 | 55.5 | 55.5 | 54.5 | 54.5 | 52.7 | 52.8 | 53.4 | 52.1 |
| 51 | 54.4 | 56.5 | 56.5 | 55.5 | 55.5 | 53.7 | 53.9 | 54.4 | 53.1 |
| 52 | 55.4 | 57.5 | 57.5 | 56.5 | 56.5 | 54.7 | 54.9 | 55.4 | 54.1 |
| 53 | 56.4 | 58.4 | 58.5 | 57.5 | 57.5 | 55.7 | 55.9 | 56.4 | 55.1 |
| 54 | 57.3 | 59.3 | 59.4 | 58.4 | 58.4 | 56.6 | 56.8 | 57.3 | 56.1 |
| 55 | 58.3 | 60.3 | 60.3 | 59.4 | 59.4 | 57.6 | 57.8 | 58.3 | 57.1 |
| 56 | 59.3 | 61.3 | 61.3 | 60.4 | 60.4 | 58.6 | 58.8 | 59.3 | 58.1 |
| 57 | 60.3 | 62.2 | 62.2 | 61.3 | 61.3 | 59.6 | 59.7 | 60.2 | 59.0 |
| 58 | 61.2 | 63.1 | 63.1 | 62.2 | 62.2 | 60.5 | 60.7 | 61.2 | 60.0 |
| 59 | 62.2 | 64.0 | 64.1 | 63.2 | 63.2 | 61.5 | 61.6 | 62.1 | 61.0 |
| 60 | 63.1 | 64.9 | 65.0 | 64.1 | 64.1 | 62.4 | 62.6 | 63.0 | 61.9 |
| 61 | 64.1 | 65.9 | 66.0 | 65.1 | 65.1 | 63.5 | 63.6 | 64.1 | 62.9 |
| 62 | 65.0 | 66.8 | 66.9 | 66.0 | 66.0 | 64.4 | 64.6 | 65.0 | 63.9 |
| 63 | 66.0 | 67.7 | 67.8 | 66.9 | 66.9 | 65.3 | 65.5 | 65.9 | 64.8 |
| 64 | 67.0 | 68.7 | 68.7 | 67.9 | 67.9 | 66.3 | 66.5 | 66.9 | 65.8 |
| 65 | 67.9 | 69.6 | 69.7 | 68.9 | 68.9 | 67.3 | 67.5 | 67.9 | 66.9 |
| 66 | 68.8 | 70.5 | 70.5 | 69.7 | 69.7 | 68.2 | 68.4 | 68.8 | 67.8 |
| 67 | 69.8 | 71.4 | 71.4 | 70.7 | 70.7 | 69.2 | 69.3 | 69.7 | 68.7 |
| 68 | 70.8 | 72.4 | 72.4 | 71.6 | 71.6 | 70.2 | 70.3 | 70.7 | 69.8 |
| 69 | 71.7 | 73.2 | 73.3 | 72.5 | 72.5 | 71.1 | 71.3 | 71.7 | 70.7 |
| 70 | 72.6 | 74.1 | 74.1 | 73.4 | 73.4 | 72.1 | 72.2 | 72.6 | 71.7 |
| 71 | 73.5 | 75 | 75.0 | 74.3 | 74.3 | 73.0 | 73.1 | 73.5 | 72.6 |
| 72 | 74.5 | 75.9 | 75.9 | 75.3 | 75.3 | 74.0 | 74.1 | 74.5 | 73.6 |
| 73 | 75.4 | 76.8 | 76.8 | 76.1 | 76.1 | 74.9 | 75.0 | 75.4 | 74.5 |
| 74 | 76.4 | 77.7 | 77.7 | 77.1 | 77.1 | 75.9 | 76.0 | 76.3 | 75.5 |
| 75 | 77.3 | 78.5 | 78.6 | 78.0 | 78.0 | 76.8 | 76.9 | 77.2 | 76.4 |
| 76 | 78.3 | 79.5 | 79.5 | 78.9 | 78.9 | 77.8 | 77.9 | 78.2 | 77.5 |
| 77 | 79.2 | 80.4 | 80.4 | 79.8 | 79.8 | 78.7 | 78.8 | 79.2 | 78.4 |
| 78 | 80.1 | 81.2 | 81.2 | 80.7 | 80.7 | 79.7 | 79.8 | 80.1 | 79.3 |
| 79 | 81.1 | 82.1 | 82.2 | 81.7 | 81.7 | 80.6 | 80.8 | 81.0 | 80.3 |
| 80 | 81.9 | 82.9 | 83.0 | 82.5 | 82.5 | 81.5 | 81.6 | 81.9 | 81.2 |
| 81 | 82.9 | 83.9 | 83.9 | 83.5 | 83.5 | 82.5 | 82.6 | 82.9 | 82.2 |
| 82 | 83.8 | 84.8 | 84.8 | 84.4 | 84.4 | 83.5 | 83.6 | 83.8 | 83.2 |
| 83 | 84.7 | 85.6 | 85.6 | 85.2 | 85.2 | 84.3 | 84.4 | 84.7 | 84.1 |
| 84 | 85.7 | 86.5 | 86.5 | 86.1 | 86.1 | 85.3 | 85.4 | 85.6 | 85.1 |
| 85 | 86.6 | 87.4 | 87.4 | 87.0 | 87.0 | 86.3 | 86.3 | 86.5 | 86.0 |
| 86 | 87.5 | 88.3 | 88.3 | 88.0 | 88.0 | 87.2 | 87.3 | 87.5 | 87.0 |
| 87 | 88.4 | 89.1 | 89.1 | 88.8 | 88.8 | 88.1 | 88.2 | 88.4 | 87.9 |
| 88 | 89.3 | 90.0 | 90.0 | 89.7 | 89.7 | 89.1 | 89.1 | 89.3 | 88.9 |
| 89 | 90.3 | 91.0 | 91.0 | 90.7 | 90.7 | 90.1 | 90.2 | 90.3 | 89.9 |
| 90 | 91.3 | 91.9 | 91.9 | 91.6 | 91.6 | 91.0 | 91.1 | 91.3 | 90.9 |
| 91 | 92.1 | 92.7 | 92.7 | 92.5 | 92.5 | 91.9 | 92.0 | 92.1 | 91.7 |
| 92 | 93.2 | 93.7 | 93.7 | 93.5 | 93.5 | 93.0 | 93.0 | 93.2 | 92.8 |
| 93 | 94.1 | 94.6 | 94.6 | 94.3 | 94.3 | 93.8 | 93.9 | 94.0 | 93.7 |
| 94 | 95.0 | 95.5 | 95.5 | 95.3 | 95.3 | 94.8 | 94.9 | 95.0 | 94.7 |
| 95 | 96.0 | 96.4 | 96.4 | 96.2 | 96.2 | 95.8 | 95.9 | 96 | 95.7 |
| 96 | 96.9 | 97.4 | 97.4 | 97.2 | 97.2 | 96.8 | 96.8 | 96.9 | 96.6 |
| 97 | 97.9 | 98.3 | 98.3 | 98.1 | 98.1 | 97.7 | 97.8 | 97.9 | 97.6 |
| 98 | 98.8 | 99.2 | 99.2 | 99.0 | 99.0 | 98.6 | 98.7 | 98.8 | 98.5 |
| 99 | 99.8 | 100.2 | 100.2 | 100.1 | 100.1 | 99.7 | 99.7 | 99.8 | 99.5 |
| 100 | 100.8 | 101.2 | 101.2 | 101.0 | 101.0 | 100.6 | 100.7 | 100.8 | 100.5 |

**Table S3:** Skeletal age for a 60-year-old individual who sustained a fracture at a specific bone.

| Fracture | Skeletal age in years (95% CI) |  |
| --- | --- | --- |
|  | Men | Women |
| Any fragility fracture | 63.8 (63.7, 63.9) | 63.1 (63.0, 63.2) |
| Specific fracture site |  |  |
| Hip | 66.1 (65.9, 66.2) | 64.9 (64.8, 65.0) |
| Femur | 65.7 (65.2, 66.1) | 64.9 (64.6, 65.2) |
| Pelvis | 65.2 (64.6, 65.8) | 64.1 (63.8, 64.4) |
| Vertebrae | 64.5 (64.2, 64.7) | 64.1 (63.9, 64.3) |
| Humerus | 64.5 (64.2, 64.8) | 62.4 (62.3, 62.6) |
| Rib | 62.6 (62.2, 63.1) | 62.6 (62.2, 63.0) |
| Clavicle | 63.3 (62.9, 63.7) | 63.0 (62.6, 63.4) |
| Lower leg | 62.4 (61.8, 62.8) | 61.9 (61.6, 62.2) |

**Figure S1:**
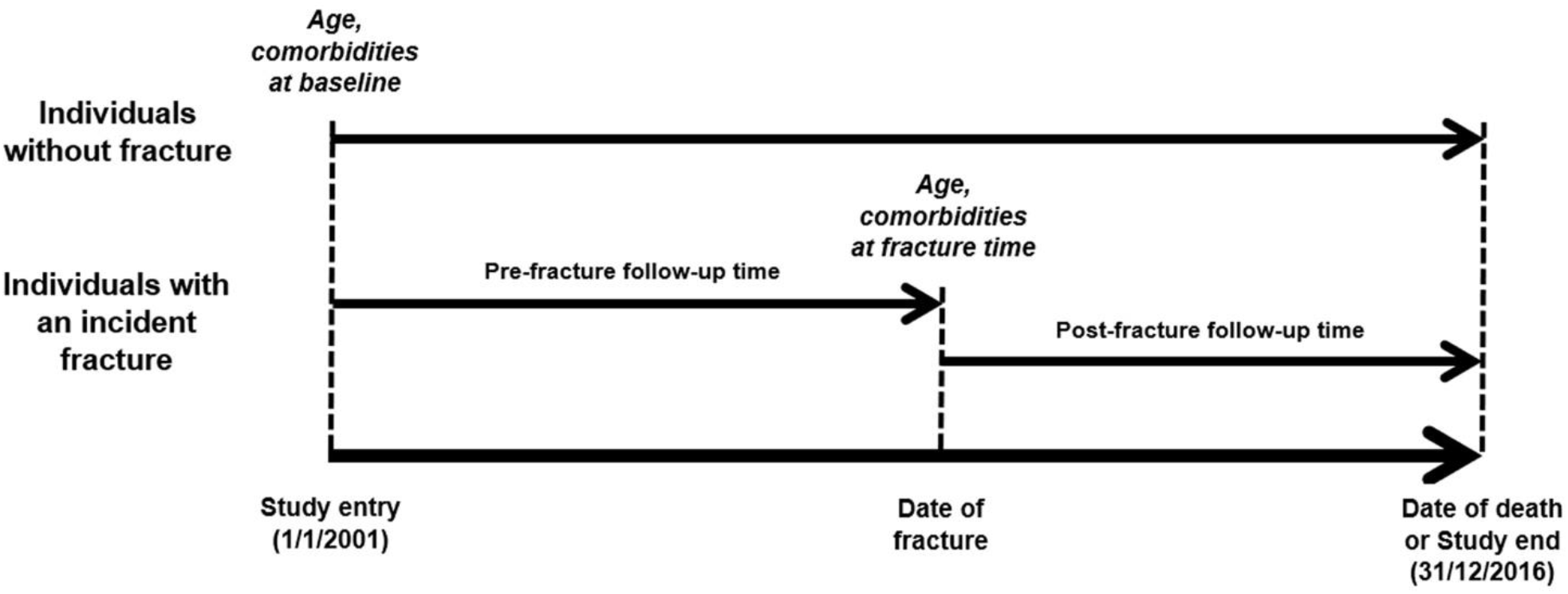
Schematic representation of time-dependent analysis.

